# MicroRNAs in adipocyte-derived extracellular vesicles in maternal and cord blood are related to neonatal adiposity

**DOI:** 10.1101/2022.10.30.22281708

**Authors:** Pooja Kunte, Matthew Barberio, Pradeep Tiwari, Krishna Sukla, Brennan Harmon, Samuel Epstein, Dattatray Bhat, Kayla Authelet, Madeleine Goldberg, Sudha Rao, Hemant Damle, Robert J Freishtat, Chittaranjan Yajnik

**Affiliations:** Diabetes Unit, KEM Hospital Research Centre, Pune, India; Department of Exercise and Nutrition Sciences, The Milken Institute School of Public Health, George Washington University, Washington, D.C.; Center for Genetic Medicine Research, Children’s National Hospital, Washington, D.C.; Genotypic Technology Pvt. Ltd., Bangalore, India; S K Navle hospital, Pune, India

**Keywords:** Adipocyte-derived extracellular vesicles (ADsEV), microRNA, maternal and cord blood, neonatal adiposity

## Abstract

**Background:** Maternal body size, nutrition, and hyperglycemia contribute to neonatal body size and composition. There is little information on maternal-fetal transmission of messages which influence fetal growth. We analyzed adipocyte-derived small extracellular vesicular (ADsEV) microRNAs in maternal and cord blood to explore their adipogenic potential.

**Methods:** We studied 127 mother-neonate pairs (51 lean and 76 adipose neonates, in 68 NGT and 59 GDM pregnancies). Adiposity refers to highest tertile (T3) of sum of skinfolds in neonates of normal glucose tolerant (NGT) mothers, lean to lowest tertile (T1). ADsEV miRNAs from maternal and cord blood samples were profiled on Agilent 8*60K microarray. Differential expression (DE) of ADsEV miRNAs in adipose vs. lean neonates was studied before and after adjustment for maternal gestational diabetes mellitus (GDM), adiposity, and vitamin B12-folate status.

**Results:** Multiple miRNAs were common in maternal and cord blood and positively correlated. We identified 24 maternal and 5 cord blood miRNAs differentially expressed (p≤0.1) in the adipose neonate group, and 19 and 26 respectively, in the adjusted analyses. Even though DE miRNAs were different in maternal and cord blood, they targeted similar adipogenic pathways (e.g., the forkhead box O (FOXO) family of transcription factors, mitogen-activated protein kinase (MAPK) pathway, transforming growth factor beta (TGF-β) pathway). Maternal GDM and adiposity were associated with many DE ADsEV miRNAs.

**Conclusion:** Our results suggest that the DE ADsEV miRNAs in mothers of adipose neonates are potential regulators of fetal adiposity. Further, the composition and functionality of miRNAs may be influenced by maternal hyperglycemia, adiposity, and micronutrient status during pregnancy.

## Introduction

Women undergo many physiological adaptations during the gestational period to promote healthy fetal growth^1^. The developing fetus depends on transfer of nutrients from the mother for the growth of its developing tissues and organs. Exposure of the fetus to overnutrition (maternal hyperglycemia) or undernutrition (maternal macro- and micro-nutrient deficiencies) during the intra-uterine development could create a ‘memory’ of malnourishment which is proposed to “program” its risk for future adiposity, diabetes, and cardiovascular diseases^2^. This is the science of the Developmental Origins of Health and Disease (DOHaD)^3^.

The Pune birth cohort studies (Pune Children’s Study (PCS) and Pune Maternal Nutrition Study (PMNS)) provided significant insight into the fetal origins of elevated risk for chronic disease^4^. The PMNS described the unique ‘thin-fat’ Indian baby which has lower birth weight and lean mass, but comparable fat mass, to a European baby^5^. The thin-fat Indian baby also has higher leptin and insulin but lower adiponectin concentrations in the cord blood^6^. This neonatal ‘thin-fat’ phenotype is thought to originate in multi-generational maternal undernutrition and is aggravated by maternal glycemia^7^. This phenotype continues into postnatal life and contributes to the higher cardiometabolic risk in Indians compared to Europeans^8^. In the Pune studies, low birth weight was associated with childhood insulin resistance, and a maternal imbalance of low vitamin B12 and high folate status was associated with high adiposity and insulin resistance in the offspring^9,10^. Prediabetes in 18-year-old offspring was predicted by poor linear growth *in utero* and by high normal glycemia in the mother during pregnancy.

Understanding the molecular pathways leading to fetal adiposity are of interest to the ‘primordial’ prevention of diabetes and other cardiometabolic diseases. Several mechanisms, such as genetics, epigenetics, nutrients, and pro-inflammatory cytokines are potential drivers of this intergenerational influence^11^. Individually, they may explain only a small proportion of the effect. For instance, genetic markers from genome wide association studies account for less than 15% of the overall observed variance in the obesity phenotypes^12,13^. More recently small extracellular vesicles (sEV), sometimes called exosomes, came into focus as potential mediators of intergenerational transfer due to their ability to cross the placenta, the rise in sEV number in the maternal circulation during pregnancy and their important role in cell-to-cell communication ^14,15^.

sEVs are endocytic vesicles with a lipid bilayer, are released by a wide range of cell types and can transfer molecular cargo (e.g., proteins, nucleic acids, lipids) from the original secreting cell to a recipient cell in a paracrine or endocrine fashion^16^. More specifically, adipocyte-derived sEVs (ADsEV) have been shown to be a major source of circulating microRNAs (miRNAs) which can regulate gene expression in distant tissues^17,18^. We have previously demonstrated that ADsEV miRNA contents are altered by obesity and normalized following significant weight-loss (i.e., from bariatric surgery)^19,20,21^. Further, ADsEVs from individuals with obesity alter macrophage cholesterol efflux and gene expression in vitro^22^. Given the important role of ADsEVs in obesity-related co-morbidities^23,24^, our objective was to explore ADsEV miRNAs as potential mediators of maternal-to-fetus communication and transfer of intergenerational risk for adiposity. In this analysis, we examined differences in miRNA profiles of ADsEVs isolated from maternal and cord blood plasma samples collected at delivery in pregnancies with adipose and lean neonates, with the hypothesis that ADsEV miRNAs will enhance adipogenic pathways.

## Methods

### Cohort description

The ‘Intergenerational programming of diabesity in offspring of women with gestational diabetes mellitus’ (InDiaGDM) study investigated the impact of maternal hyperglycemia on the risk of diabesity (diabetes + adiposity) in the offspring and the modifying influence of vitamin B12 and folate status^25^. Details of inclusion and exclusion criteria are shown in the flowchart (Supplementary figure 1). All participating women signed an informed consent, and the study was approved by the institutional ethics committee (KEM EC no. 1404, SKNMC /Ethics/App/2014/265).

#### Maternal and Neonatal Data

*M*aternal socio-demographic data, medical and obstetric information, and standard anthropometric measurements were taken at 28 weeks gestation. A fasting oral glucose tolerance test (OGTT, 75-gram anhydrous glucose, IADPSG^26^) was performed at that time. Those diagnosed with gestational diabetes mellitus (GDM) were appropriately advised. Delivery details (gestational age and type of delivery (e.g., vaginal, cesarean)) were recorded. Detailed neonatal anthropometry was performed within 24 hours of delivery (i.e., weight, length, head, and abdominal circumferences, triceps, and subscapular skin folds).

### Biospecimen collection and determination of blood metabolites

The following measurements were made on maternal and cord blood: hemogram (complete blood count), plasma glucose, insulin, total and HDL cholesterol, triglycerides, total vitamin B12, total homocysteine and folate. Supplementary table 1 describes laboratory methods used for these measurements.

### Definitions

Neonatal adiposity (sex-stratified) was defined as highest tertile (T3) of sum of skinfolds (SSF) of neonates born to normal glucose tolerant (NGT) mothers. T1 were classified as lean. Maternal adiposity was defined from sub-scapular skin folds at 28 weeks’ gestation: above median (adipose) and below median (lean). GDM was diagnosed using IADPSG criteria^26^. Maternal vitamin B12 and folate status were classified as adequate if above and inadequate if below the median concentration for the cohort.

### Sample processing randomization and blinding

To avoid unintentional sampling bias, the sample IDs were randomly ordered five times and then mother-neonate pairs were grouped into batches of eight pairs (N=16). Laboratory staff were provided with a blinded (i.e., no group information or metadata) processing order for samples.

### ADsEV isolation

Maternal and cord blood plasma samples at delivery were treated with 10μL [611U/mL] Thrombin (System Biosciences, Palo Alto, CA, USA) per 0.5mL plasma. After a 10-minute incubation at room temperature, and centrifugation in a standard microcentrifuge at 12,298 g for 10 minutes, the supernatant was removed and filtered through a 0.2μm cellulose acetate syringe filter (VWR International, Radnor, PA, USA). Total EVs were isolated from filtered plasma supernatants using ExoQuick exosome precipitation solution (System Biosciences, Palo Alto, CA, USA) according to manufacturer’s instructions. ADsEVs were positively selected with a fatty acid-binding protein antibody (FABP4) (0.5 mg/mL; Mouse IgG monoclonal antibody, Abgent cat. No AM2235b, Inc., San Diego, CA, USA) using a magnetic capture technique (EasySep™ “Do-It-Yourself” Positive Selection Kit II - StemCell Technologies, Vancouver, BC, Canada). After positive selection, the ADsEVs were resuspended in TRIzol™ Reagent (ThermoFisher Scientific, Waltham, MA, USA).

### MicroRNA isolation and assessment

Exosomal RNA isolation was completed using TRIzol reagent (ThermoFisher Scientific, Waltham, MA USA) according to manufacturer’s instructions. The quantity and quality of the RNA were assessed using a Nanodrop 2000 (ThermoFisher Scientific, Waltham, MA USA) and Bioanalyzer 2100 (Agilent Technologies, Santa Clara, CA USA). Samples with OD260/OD280 ratio >1.5 and <2.2 were selected for miRNA profiling. The miRNA labeling was performed using miRNA Complete Labeling and Hyb Kit (Agilent Technologies, Santa Clara, CA, USA) with a sample input of 10uL of total RNA. Samples were hybridized to Agilent Human miRNA Microarray, Release 21.0, 8×60K. The microarray slides were scanned using an Agilent scanner (G2600D) and raw data extracted using Agilent Feature Extraction software (Version 11.5.1.1).

### Statistical analyses

Sample size and power calculations were done using “ssize” package in R to measure a reliable fold change of 2.0 in miRNA expression in the adipose group with respect to lean at a significance level of 0.05. A total of 120 individuals (∼60 in each group) would provide a power of 80%.

### Demographic characteristics

Normally distributed data are presented as mean (SD) and skewed data as median (25^th^ and 75^th^ percentiles). The significance of differences between groups was tested by Student’s t test (if normally distributed) or Mann-Whitney test (if skewed).

### MicroRNA data

Raw microRNA expression data was log2 transformed and normalized using the 90th Percentile shift normalization method in GeneSpring GX (Version 14.5) software. MiRNAs that showed true signal in at least 10% of samples were selected for further analysis. To check if the average expression of all ADsEV miRNAs was upregulated or downregulated in the adipose group, mean z-scores of miRNA expression values were calculated for all miRNAs from lean and adipose groups in maternal plasma and cord blood and the distribution of z-scores between the groups were compared. We also investigated if the ADsEV miRNA expression in maternal plasma samples correlated with those from cord blood samples by applying a Spearman correlation test.

The central objective of the study was to compare differential expression of ADsEV miRNAs in maternal plasma and cord blood of adipose (T3) and lean neonates (T1). Differential expression of miRNAs between groups was calculated by Student’s T test for those normally distributed and by Mann-Whitney test for skewed. Fold changes were calculated by formula: *FC = average (miRNA expression in adipose – median expression of miRNA in lean)*. This was done in 2 steps: 1. Without adjusting for confounding variables and 2. with adjusting for confounding variables (maternal GDM status, adiposity, and maternal vitamin B12 and folate status for maternal samples, and additionally neonatal gender for cord blood samples) by multiple linear regression analysis. We used a discovery significance of p≤0.1 and a FC≥|1.2| and represented it as a volcano plot generated using ggplot2^25^ package in R.

### Functional analyses

Differentially expressed miRNAs with p-value≤0.1 were selected for miRNA target gene and functional enrichment using Mienturnet^27^ software (http://userver.bio.uniroma1.it/apps/mienturnet/; March 2019). Target enrichment was done using miRTarBase 8.0 and the functional enrichment was conducted by using the KEGG database. This tool computationally maps the given list of miRNAs to their target genes/proteins in their reference database (KEGG/miRTarBase) and fetches a list of target genes that are computationally predicted and/or experimentally confirmed. Relevant overrepresented classes of annotated genes with at least 1 miRNA-target interaction were selected for functional enrichment and are studied. MicroRNAs targeting biological pathways belonging to the following KEGG pathway categories are not shown because they are of no direct interest in this analysis: cancer, bacterial/viral/parasitic infectious diseases, excretory, immune and nervous system, immune and neurodegenerative diseases, environmental adaptation, aging, substance abuse, and drug resistance. Pathways with FDR p-value≤0.05 and related to adipogenesis, adipocyte differentiation and adipocyte signaling were considered relevant from the KEGG pathway categories like cell growth and development, signal transduction, metabolism, endocrine and cardiovascular health, digestive systems and studied in detail. Fold changes of each DE miRNA in adipose group are shown in a barplot. MiRNAs and their targeted genes are represented as a network plot, generated using ggnet^28^ package in R. A bubble plot of the functionally enriched miRNAs was generated using ggplot2^29^, where colour and size of each bubble stands for its FDR p-value and the number of targeted genes, respectively. Adipogenesis related pathways targeted by both maternal and cord blood DE miRNAs are highlighted using a Sankey diagram generated using SankeyMatic^30^ software.

### Differential expression of miRNAs in relation to maternal conditions (hyperglycemia and adiposity during pregnancy)

To check the influence of maternal conditions (i.e., hyperglycemia and adiposity during pregnancy) on ADsEV miRNA profile, differential expression analysis was carried out in both maternal and cord blood ADsEV miRNA profiles between following groups: 1. GDM and NGT (unadjusted and adjusted for confounders: maternal adiposity, B12 and folate status) and 2. maternal adiposity (below and above median) (unadjusted and adjusted for confounders: GDM, B12 and folate status). The lists of DE miRNAs from the unadjusted and adjusted analyses were combined and duplicates were removed. DE miRNAs from both analyses were subjected to functional enrichment in Mienturnet and adipogenesis related pathways were screened. A Venn diagram was made using jvenn^31^ software to compare the adipogenesis related microRNAs from the three analyses: 1. Neonatal adiposity, 2. Maternal GDM, and 3. Maternal adiposity in both maternal and cord blood.

## Results

The InDiaGDM study enrolled 413 women after applying inclusion and exclusion criteria. Of these, 330 women delivered at study centers. Two hundred and seventy-nine mother-neonate pairs had the necessary data and matching maternal plasma and cord blood samples. From these, guided by a power calculation, we randomly selected 127 mother-neonate pairs, including 51 lean and 76 adipose neonates, for ADsEV miRNA analyses (supplementary figure 1). There were no significant differences between the included (N=127) and excluded (N=152) mothers with respect to age, parity, weight, and BMI, or in their neonates with respect to gender, gestational age at the time of birth, and birth weight (supplementary table 2).

### Characteristics of mothers and neonates

Mothers were a median (IQR) age of 27 (5) years old, 155 (7) cm tall, and had a BMI of 27.1 (6.6) kg/m^2^ at 28 weeks’ gestation. Of the 127 mothers, 68 were NGT and 59 were diagnosed with GDM. The mothers of adipose neonates had higher weight, BMI, SSF, and fasting glucose compared to mothers of lean neonates but similar levels of total and HDL cholesterol, triglycerides, vitamin B12, and folate (Table 1A). Adipose neonates had higher birth weight, length, SSF, and placental weight compared to lean neonates (Table 1B). Mothers of adipose neonates had higher prevalence of adiposity and GDM.

**Table 1.**
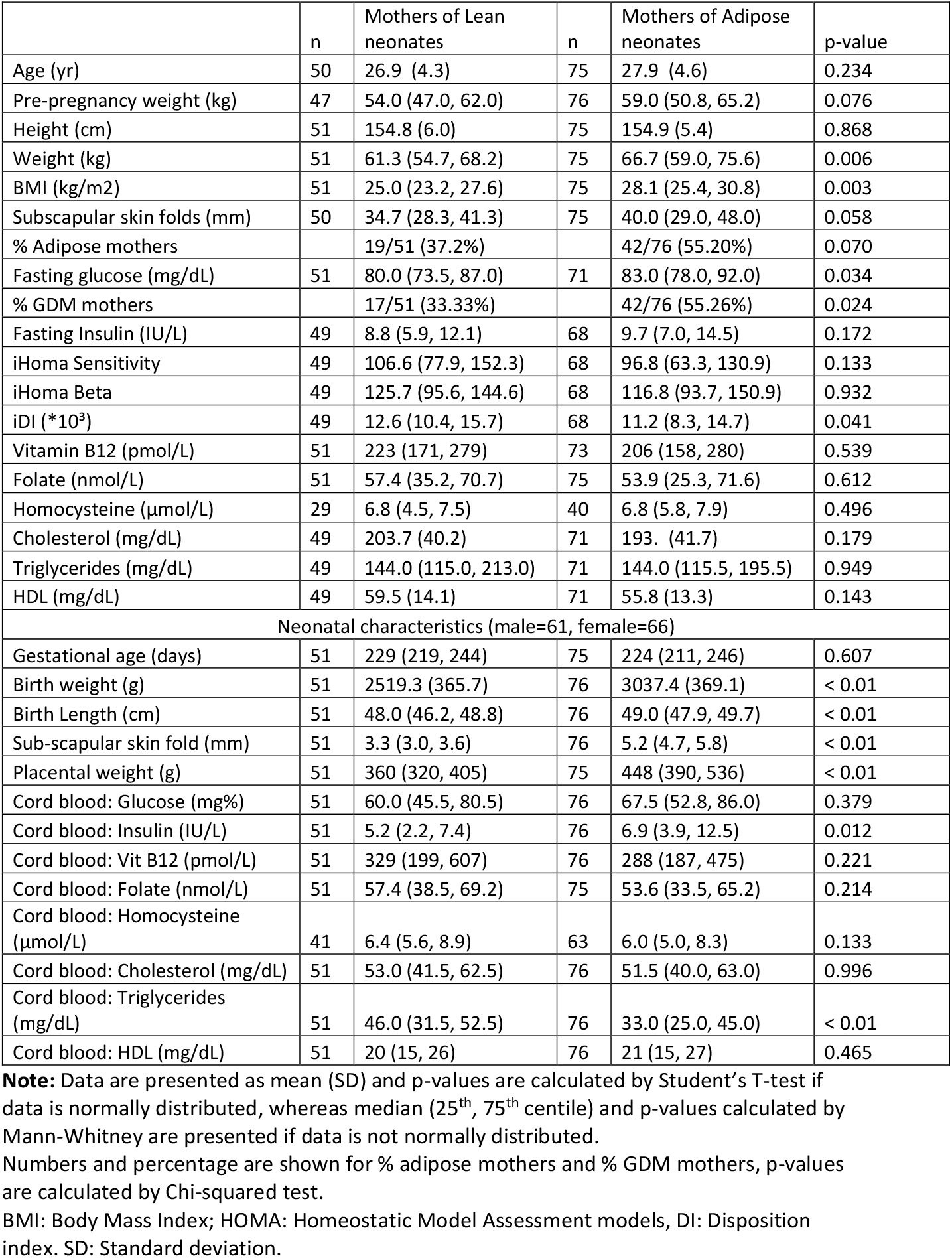
Comparison of demographic characteristics between mothers of lean vs. mothers of adipose neonates, and lean vs. adipose neonates.

|  | n | Mothers of Lean neonates | n | Mothers of Adipose neonates | p-value |
| --- | --- | --- | --- | --- | --- |
| Age (yr) | 50 | 26.9 (4.3) | 75 | 27.9 (4.6) | 0.234 |
| Pre-pregnancy weight (kg) | 47 | 54.0 (47.0, 62.0) | 76 | 59.0 (50.8, 65.2) | 0.076 |
| Height (cm) | 51 | 154.8 (6.0) | 75 | 154.9 (5.4) | 0.868 |
| Weight (kg) | 51 | 61.3 (54.7, 68.2) | 75 | 66.7 (59.0, 75.6) | 0.006 |
| BMI (kg/m<sup>2</sup>) | 51 | 25.0 (23.2, 27.6) | 75 | 28.1 (25.4, 30.8) | 0.003 |
| Subscapular skin folds (mm) | 50 | 34.7 (28.3, 41.3) | 75 | 40.0 (29.0, 48.0) | 0.058 |
| % Adipose mothers |  | 19/51 (37.2%) |  | 42/76 (55.20%) | 0.070 |
| Fasting glucose (mg/dL) | 51 | 80.0 (73.5, 87.0) | 71 | 83.0 (78.0, 92.0) | 0.034 |
| % GDM mothers |  | 17/51 (33.33%) |  | 42/76 (55.26%) | 0.024 |
| Fasting Insulin (IU/L) | 49 | 8.8 (5.9, 12.1) | 68 | 9.7 (7.0, 14.5) | 0.172 |
| iHoma Sensitivity | 49 | 106.6 (77.9, 152.3) | 68 | 96.8 (63.3, 130.9) | 0.133 |
| iHoma Beta | 49 | 125.7 (95.6, 144.6) | 68 | 116.8 (93.7, 150.9) | 0.932 |
| iDI (*10<sup>3</sup>) | 49 | 12.6 (10.4, 15.7) | 68 | 11.2 (8.3, 14.7) | 0.041 |
| Vitamin B12 (pmol/L) | 51 | 223 (171, 279) | 73 | 206 (158, 280) | 0.539 |
| Folate (nmol/L) | 51 | 57.4 (35.2, 70.7) | 75 | 53.9 (25.3, 71.6) | 0.612 |
| Homocysteine (μmol/L) | 29 | 6.8 (4.5, 7.5) | 40 | 6.8 (5.8, 7.9) | 0.496 |
| Cholesterol (mg/dL) | 49 | 203.7 (40.2) | 71 | 193. (41.7) | 0.179 |
| Triglycerides (mg/dL) | 49 | 144.0 (115.0, 213.0) | 71 | 144.0 (115.5, 195.5) | 0.949 |
| HDL (mg/dL) | 49 | 59.5 (14.1) | 71 | 55.8 (13.3) | 0.143 |
| Neonatal characteristics (male=61, female=66) |  |  |  |  |  |
| Gestational age (days) | 51 | 229 (219, 244) | 75 | 224 (211, 246) | 0.607 |
| Birth weight (g) | 51 | 2519.3 (365.7) | 76 | 3037.4 (369.1) | < 0.01 |
| Birth Length (cm) | 51 | 48.0 (46.2, 48.8) | 76 | 49.0 (47.9, 49.7) | < 0.01 |
| Sub-scapular skin fold (mm) | 51 | 3.3 (3.0, 3.6) | 76 | 5.2 (4.7, 5.8) | < 0.01 |
| Placental weight (g) | 51 | 360 (320, 405) | 75 | 448 (390, 536) | < 0.01 |
| Cord blood: Glucose (mg%) | 51 | 60.0 (45.5, 80.5) | 76 | 67.5 (52.8, 86.0) | 0.379 |
| Cord blood: Insulin (IU/L) | 51 | 5.2 (2.2, 7.4) | 76 | 6.9 (3.9, 12.5) | 0.012 |
| Cord blood: Vit B12 (pmol/L) | 51 | 329 (199, 607) | 76 | 288 (187, 475) | 0.221 |
| Cord blood: Folate (nmol/L) | 51 | 57.4 (38.5, 69.2) | 75 | 53.6 (33.5, 65.2) | 0.214 |
| Cord blood: Homocysteine (μmol/L) | 41 | 6.4 (5.6, 8.9) | 63 | 6.0 (5.0, 8.3) | 0.133 |
| Cord blood: Cholesterol (mg/dL) | 51 | 53.0 (41.5, 62.5) | 76 | 51.5 (40.0, 63.0) | 0.996 |
| Cord blood: Triglycerides (mg/dL) | 51 | 46.0 (31.5, 52.5) | 76 | 33.0 (25.0, 45.0) | < 0.01 |
| Cord blood: HDL (mg/dL) | 51 | 20 (15, 26) | 76 | 21 (15, 27) | 0.465 |

**Note:** Data are presented as mean (SD) and p-values are calculated by Student's T-test if data is normally distributed, whereas median (25<sup>th</sup>, 75<sup>th</sup> centile) and p-values calculated by Mann-Whitney are presented if data is not normally distributed.
Numbers and percentage are shown for % adipose mothers and % GDM mothers, p-values are calculated by Chi-squared test.
BMI: Body Mass Index; HOMA: Homeostatic Model Assessment models, DI: Disposition index. SD: Standard deviation.

### Differential expression of ADsEV miRNAs in maternal plasma and cord blood

Of the 2549 miRNAs profiled in the Agilent 8*60k microarray, 742 miRNAs were detected in at least 1 sample. We chose miRNAs which were expressed in at least 10% of samples for further analysis (139 miRNAs in maternal and 144 in cord blood) (supplementary figure 2). We found that 118 miRNAs were common in maternal and cord blood. Of these, 74 (63%) were positively and significantly correlated (p≤0.05).

Comparison between the adipose and lean neonate groups identified 24 miRNAs differentially expressed in their mothers at p≤0.1 (13 with FC≥1.25 and 1 with FC≤-1.25). After adjusting for maternal GDM, adiposity, vitamin B12, and folate status, 19 miRNAs remained differentially expressed between the groups (5 with FC≥1.25 and 1 with FC≤-1.25) (Table 3A, Figure 1). In the cord blood, 5 ADsEV miRNAs were differentially expressed (3 with FC≥1.25 in adipose group). After adjusting for neonatal sex, maternal GDM, adiposity, vitamin B12, and folate status, the number of DE miRNAs in the cord blood increased to 26 (16 with FC≥1.25 and 3 with FC≤-1.25 in adipose group) (Table 3B, Figure 1). After combining the lists of DE miRNAs in unadjusted and adjusted analyses and removing duplicates, 39 miRNAs in maternal and 29 miRNAs in cord blood were found to be differentially expressed in adipose group.

**Figure 1.**
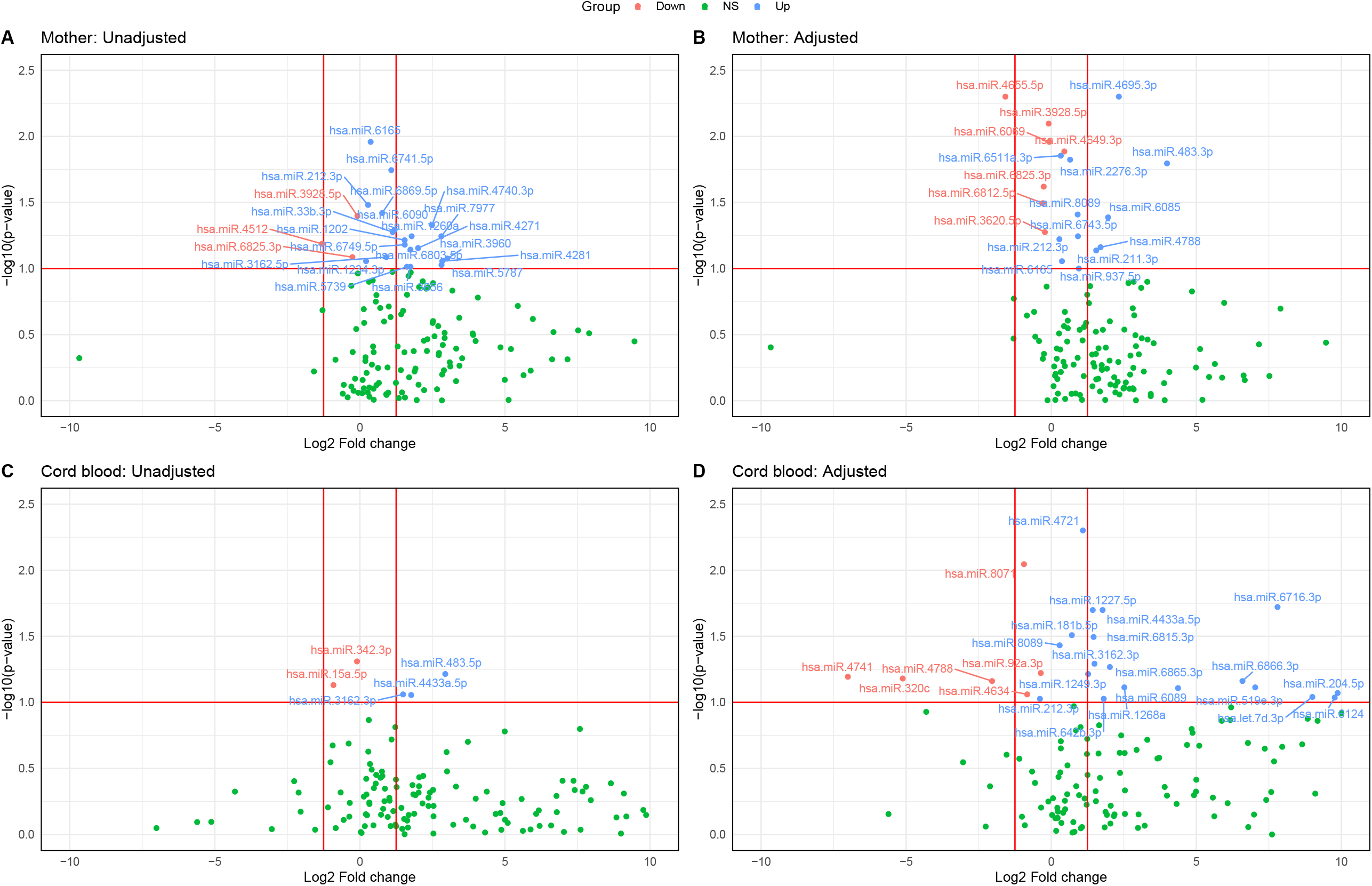
Differentially expressed miRNAs between lean Vs adipose groups in maternal and cord blood samples. Volcano plots of maternal (A, B) and cord blood (C, D) miRNAs. Significantly upregulated are shown in blue and downregulated in red, non-significant in green. A fold change cut off ±1.25 and a p-value cut-off of 0.1 are shown. Adjustment included maternal GDM, adiposity, vitamin B12 and folate status, and additionally for neonatal gender in cord blood.

We investigated biological functional profile of the differentially expressed miRNAs. Target enrichment analysis (Mienturnet) highlighted 11 miRNAs to be upregulated in plasma of mothers of adipose neonates compared to those of lean neonates (7 miRNAs in unadjusted analysis: hsa-miR-1202, hsa-miR-1234-3p, hsa-miR-212-3p, hsa-miR-33b-3p, hsa-miR-451a, hsa-miR-4271 and hsa-miR-5787; and 4 miRNAs in the adjusted analyses: hsa-miR-211-3p, hsa-miR-212-3p, hsa-miR-483-3p, hsa-miR-4649-3p). The fold changes are shown in Table 2 and Figures 2 and 3. Further screening based on KEGG ontology revealed 4 miRNAs to be targeting genes involved in pathways related to adipogenesis.

**Table 2A.** Lists of differentially expressed ADsEV miRNAs in mothers of adipose Vs lean neonates.

| Maternal miRNA (unadjusted) |  |  | Maternal miRNA (adjusted) |  |  |
| --- | --- | --- | --- | --- | --- |
| miRNA | P-value | Fold change | miRNA | P-value | Fold change |
| hsa-miR-1202 | 0.061 | 1.544 | hsa-miR-211-3p | 0.073 | 1.55 |
| hsa-miR-1234-3p | 0.088 | 0.214 | hsa-miR-212-3p | 0.06 | 0.28 |
| hsa-miR-1260a | 0.057 | 1.784 | hsa-miR-2276-3p | 0.015 | 0.65 |
| hsa-miR-212-3p | 0.033 | 0.282 | hsa-miR-3620-5p | 0.053 | -0.22 |
| hsa-miR-3162-5p | 0.082 | 0.911 | hsa-miR-3928-5p | 0.008 | -0.09 |
| hsa-miR-33b-3p | 0.053 | 1.13 | hsa-miR-4649-3p | 0.013 | 0.45 |
| hsa-miR-3656 | 0.097 | 1.744 | hsa-miR-4655-5p | 0.005 | -1.58 |
| hsa-miR-3928-5p | 0.04 | -0.085 | hsa-miR-4695-3p | 0.005 | 2.33 |
| hsa-miR-3960 | 0.084 | 3.018 | hsa-miR-4788 | 0.069 | 1.7 |
| hsa-miR-4271 | 0.07 | 2.006 | hsa-miR-483-3p | 0.016 | 3.99 |
| hsa-miR-4281 | 0.088 | 2.84 | hsa-miR-6069 | 0.011 | -0.07 |
| hsa-miR-4512 | 0.065 | -1.302 | hsa-miR-6085 | 0.041 | 1.96 |
| hsa-miR-451a | 0.099 | 40.596 | hsa-miR-6165 | 0.088 | 0.37 |
| hsa-miR-4740-3p | 0.047 | 2.488 | hsa-miR-6511a-3p | 0.014 | 0.33 |
| hsa-miR-5739 | 0.097 | 1.622 | hsa-miR-6743-5p | 0.057 | 0.92 |
| hsa-miR-5787 | 0.094 | 2.814 | hsa-miR-6812-5p | 0.032 | -0.29 |
| hsa-miR-6090 | 0.051 | 1.198 | hsa-miR-6825-3p | 0.024 | -0.26 |
| hsa-miR-6165 | 0.011 | 0.372 | hsa-miR-8089 | 0.039 | 0.91 |
| hsa-miR-6741-5p | 0.018 | 1.083 | hsa-miR-937-5p | 0.1 | 0.95 |
| hsa-miR-6749-5p | 0.066 | 1.547 |  |  |  |
| hsa-miR-6803-5p | 0.072 | 1.743 |  |  |  |
| hsa-miR-6825-3p | 0.082 | -0.257 |  |  |  |
| hsa-miR-6869-5p | 0.038 | 0.769 |  |  |  |
| hsa-miR-7977 | 0.057 | 2.809 |  |  |  |

**Note:** Lists of differentially expressed miRNAs in mothers of lean vs. adipose neonates; 24 miRNAs in the unadjusted analysis and 19 miRNAs in adjusted analysis (adjusted for maternal glycemic status (GDM/NGT), maternal adiposity (lean if below median and adipose if above median), and maternal vitamin B12 and folate status (inadequate if below median and adequate if above median)) were found to be differentially expressed at $p < 0.1$ .

**Table 2B.**
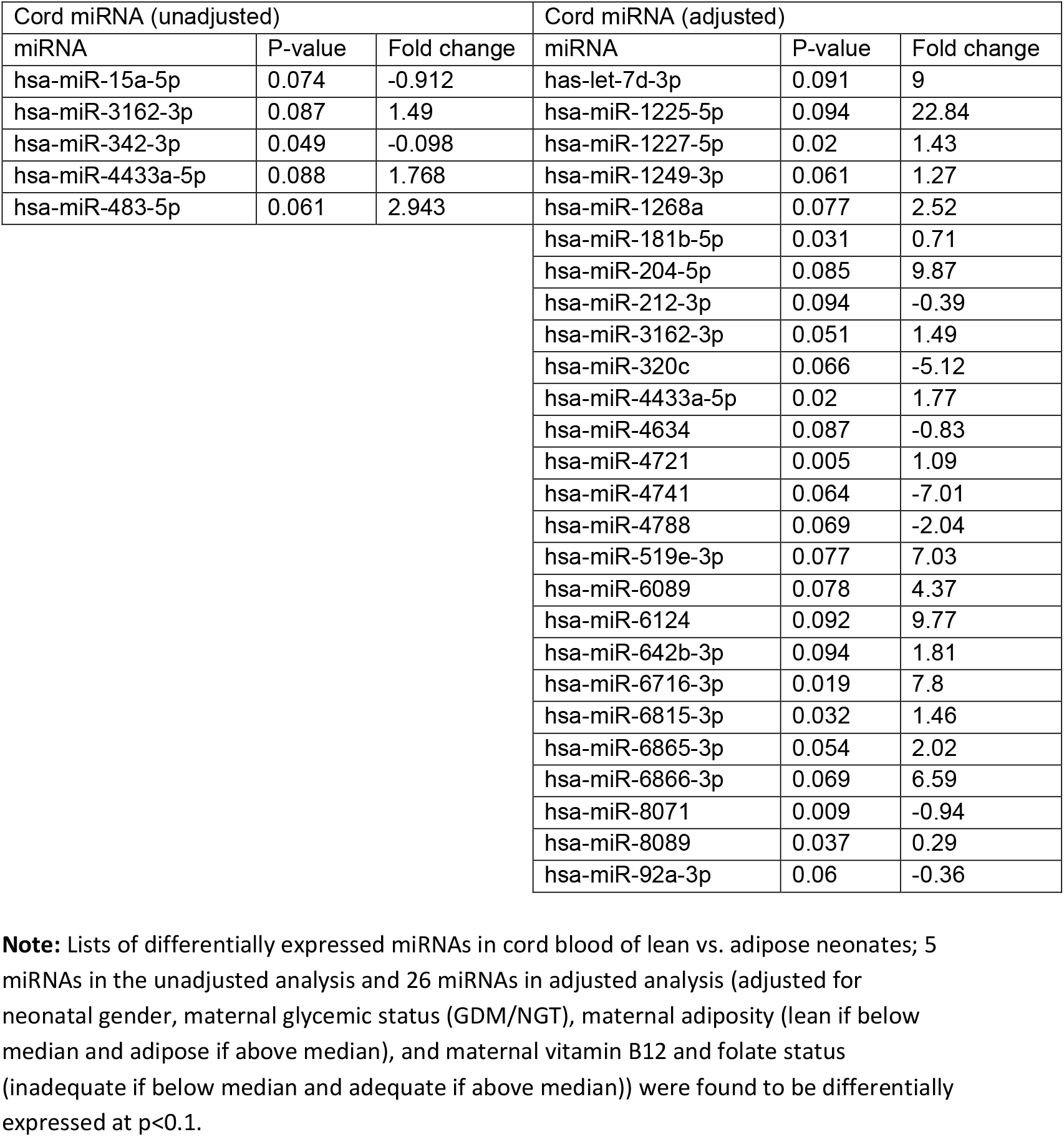
Lists of differentially expressed ADsEV miRNAs in cord blood of adipose Vs lean neonates.

**Figure 2A.**
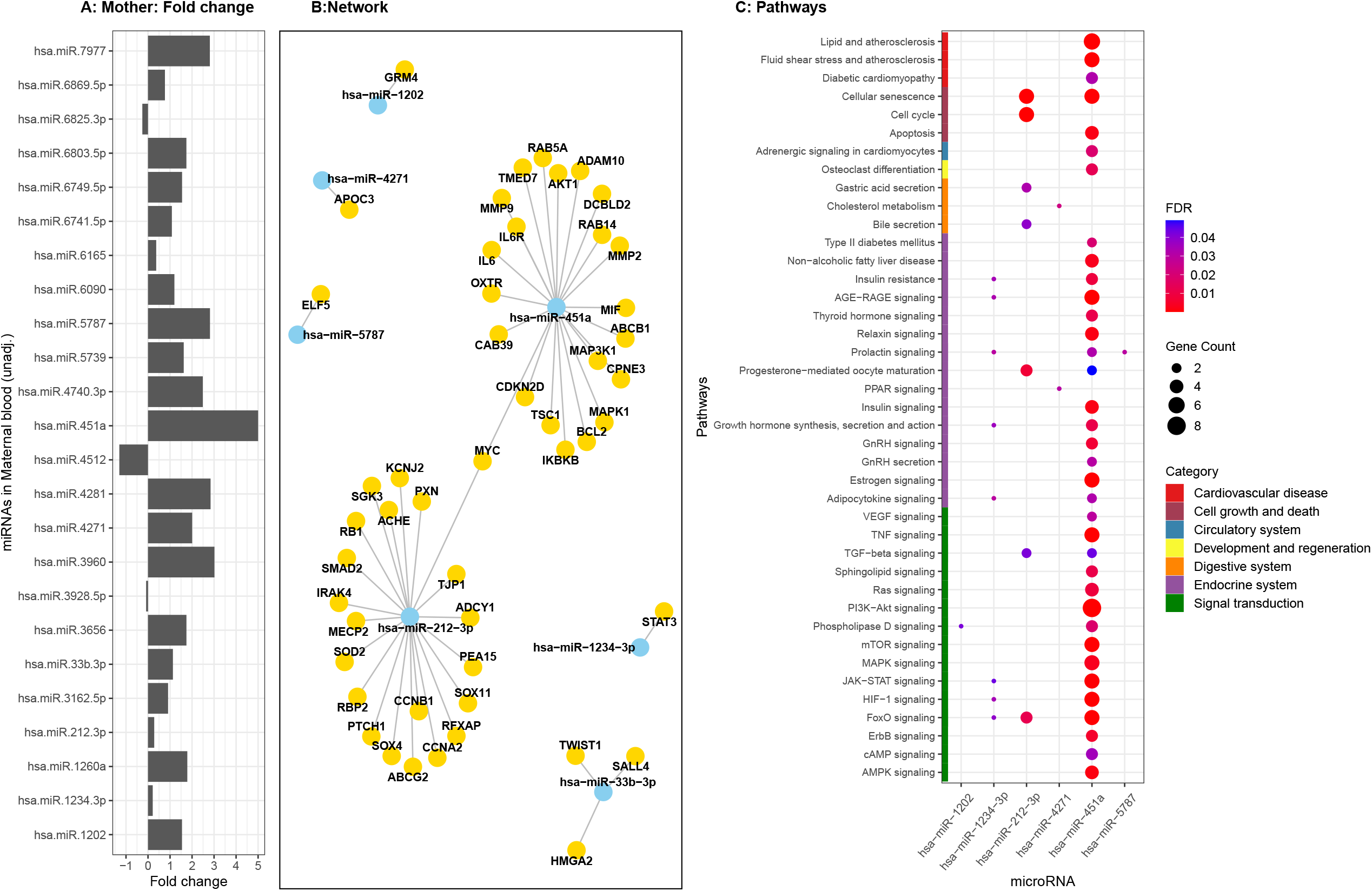
Differentially expressed ADsEV miRNAs highlighted in maternal unadjusted analysis: The bar diagram (A) shows the DE miRNAs and their fold change expression in the mothers of adipose neonates (unadjusted analysis) compared to mothers of lean neonates. Fold change values have been capped to ± 5. The network plot (B) highlights functionally enriched miRNAs from this analysis and their target genes. The bubble plot (C) reveals different adipogenesis related pathways targeted by the functionally enriched miRNAs.

**Figure 2B.**
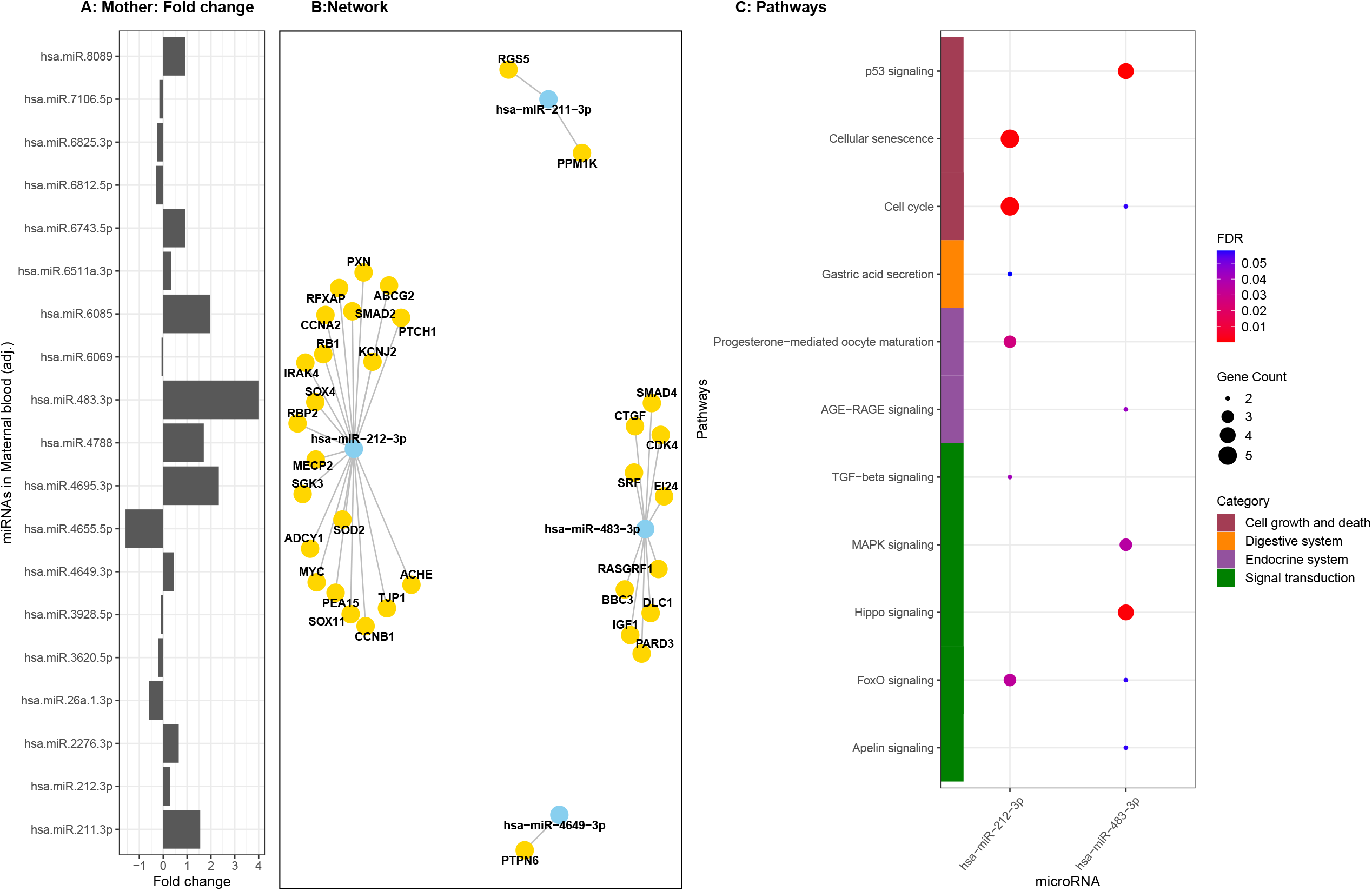
Differentially expressed ADsEV miRNAs highlighted in maternal adjusted analysis: The bar diagram (A) shows the DE miRNAs and their fold change expression in the mothers of adipose neonates compared to mothers of lean neonates (adjusted analysis; adjustments included maternal GDM, adiposity, vitamin B12 and folate status). Fold change values have been capped to ± 5. The network plot (B) highlights functionally enriched miRNAs from this analysis and their target genes. The bubble plot (C) reveals different adipogenesis related pathways targeted by the functionally enriched miRNAs.

**Figure 3A:**
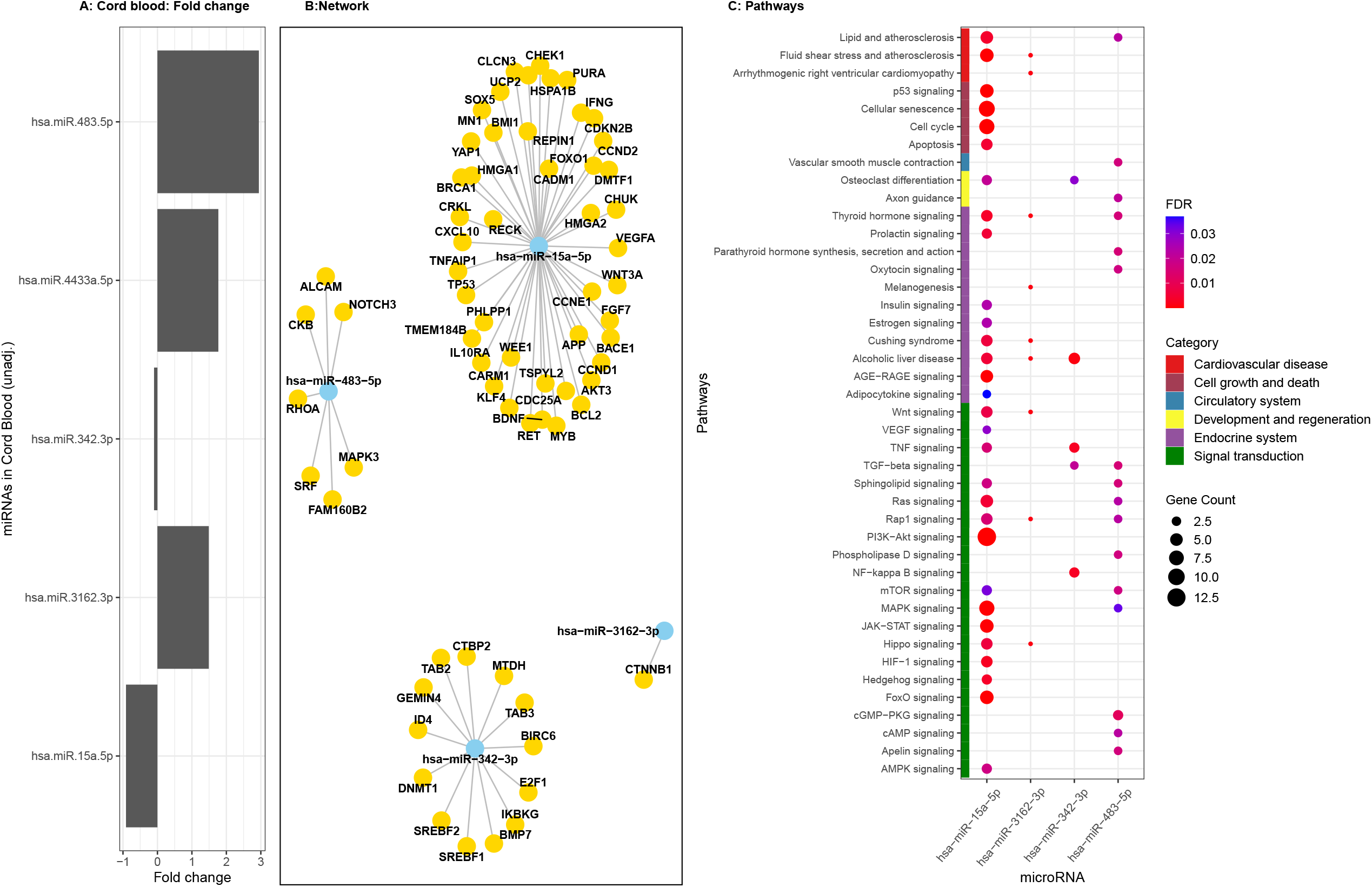
Differentially expressed ADsEV miRNAs highlighted in cord blood unadjusted analysis: The bar diagram (A) shows the DE miRNAs and their fold change expression in the cord blood of adipose neonates (unadjusted analysis) compared to lean neonates. Fold change values have been capped to ± 5. The network plot (B) highlights functionally enriched miRNAs from this analysis and their target genes. The bubble plot (C) reveals different adipogenesis-related pathways targeted by the functionally enriched miRNAs.

**Figure 3B:**
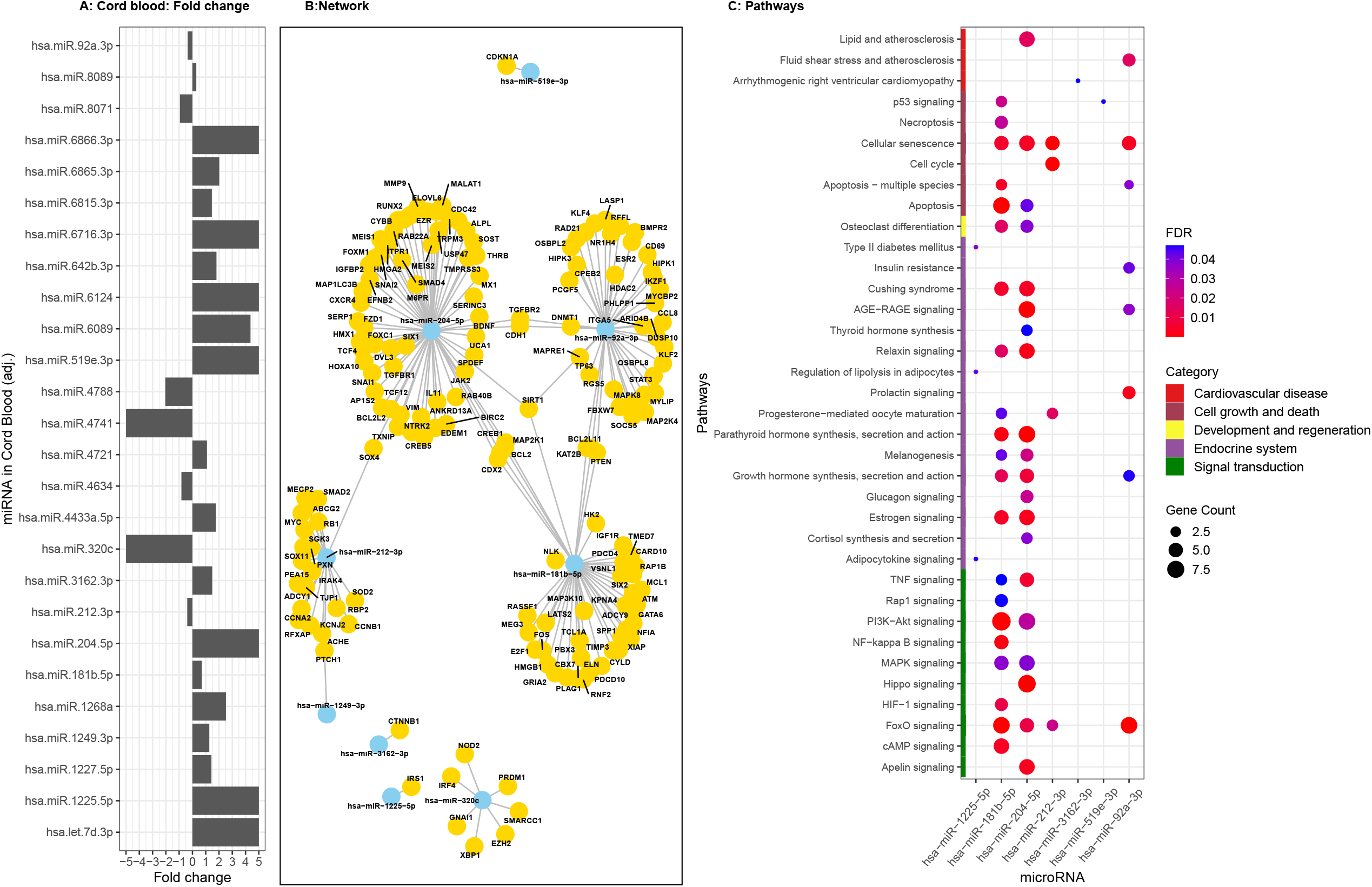
Differentially expressed ADsEV miRNAs highlighted in cord blood adjusted analysis: The bar diagram (A) shows the DE miRNAs and their fold change expression in the cord blood of adipose neonates compared to lean neonates (adjusted analysis; adjustments included maternal GDM, adiposity, vitamin B12 and folate status, and additionally for neonatal gender in cord blood). Fold change values have been capped to ± 5. The network plot (B) highlights functionally enriched miRNAs from this analysis and their target genes. The bubble plot (C) reveals different adipogenesis-related pathways targeted by the functionally enriched miRNAs.

Target enrichment analysis in the cord blood highlighted 11 miRNAs (4 miRNAs in unadjusted analysis, two upregulated: hsa-miR-483-5p, hsa-miR-3162-3p, two downregulated: hsa-miR-15a-3p, hsa-miR-342-3p; and 7 miRNAs in the adjusted analysis five upregulated: hsa-miR-1225-5p, hsa-miR-181b-5p, hsa-miR-204-5p, hsa-miR-3162-3p, hsa-miR-519e-3p; and two downregulated: hsa-miR-92a-3p, hsa-miR-212-3p). Based on KEGG ontology, 9 miRNAs of these were targeting genes involved in pathways related to adipogenesis.

We used a Sankey diagram (figure 4) to depict the inter-relationships between differentially expressed miRNAs in maternal and cord blood (of adipose neonates) and their targeted adipogenic pathways. The highlighted pathways included adipocytokine signaling, FoxO signaling, MAPK signaling, Hedgehog signaling, Wnt signaling, insulin signaling, AGE-RAGE signaling, cAMP signaling, JAK-STAT signaling, Ras signaling, and TGF-beta signaling.

**Figure 4:**
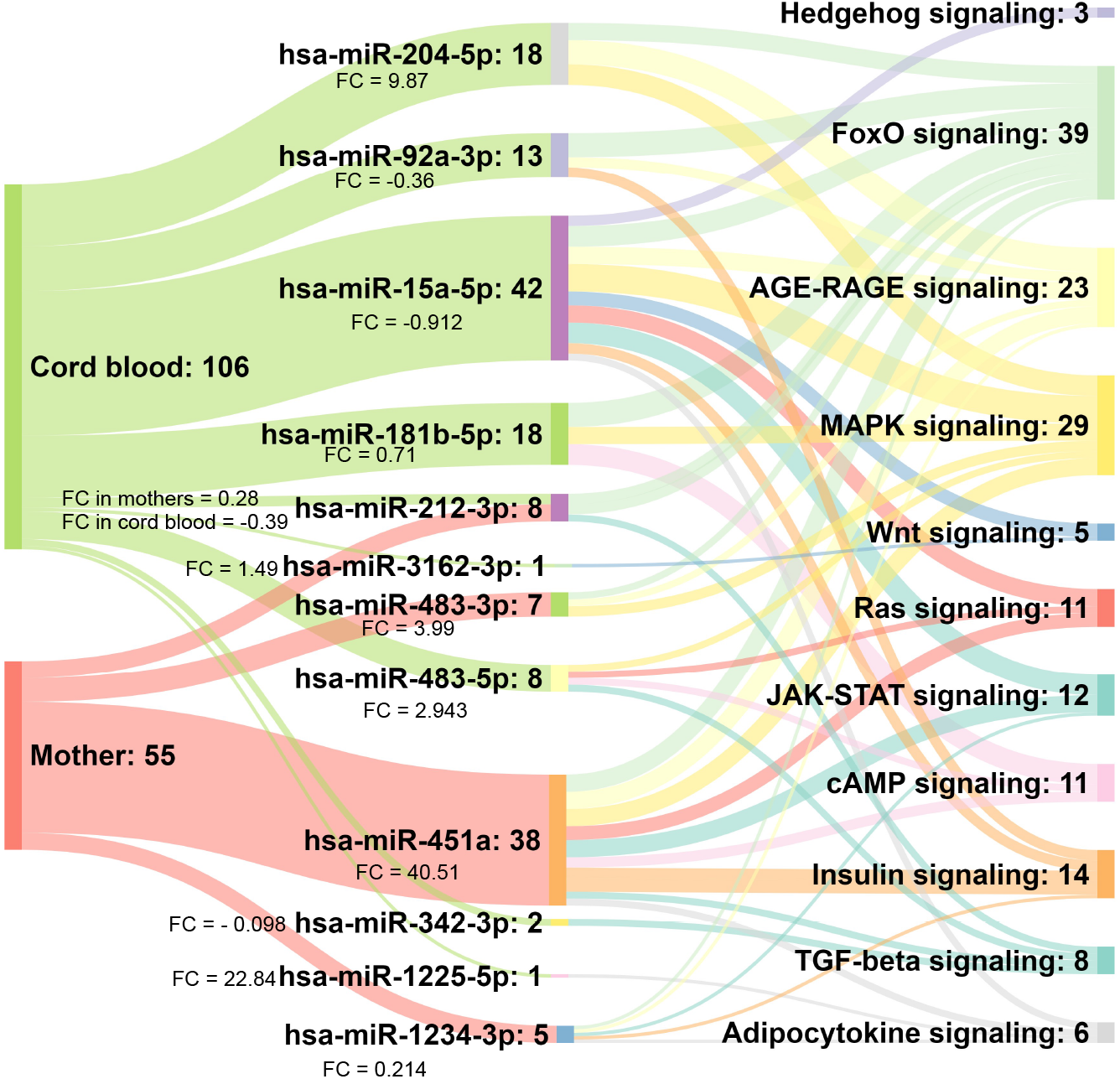
Complex interplay of DE ADsEVs miRNAs from maternal and cord blood in adipogenesis related pathways. The Sankey diagram shows the DE miRNAs in maternal and cord blood of adipose neonates and their targeted pathways related to adipogenesis. The diagram highlights that though the differentially expressed miRNAs were different in maternal and cord blood, they targeted similar adipogenesis-related pathways. The numbers represent the total number of targeted genes.

We compared differential expression of ADsEV miRNAs in plasma of adipose vs. non-adipose and in GDM vs NGT mothers and in the cord blood of their neonates. We found 127 DE ADsEV miRNAs in adipose and 106 in GDM mothers, and 57 and 82 in the cord blood, respectively. Of these, 25 miRNAs in the adipose and 19 in the GDM mothers were related to adipogenesis, the corresponding number in the cord blood was 16 and 17 respectively.

Figure 5 shows that 28 miRNAs were common in maternal blood amongst adipose mothers, GDM mothers and adipose neonates and 3 of them were related to adipogenesis: hsa-miR-451a, hsa-miR-212-3p, hsa-miR-1234-3p and 13 miRNAs were common in cord blood of which only 1 hsa-miR-204-5p was related to adipogenesis based on KEGG ontology.

**Figure 5:**
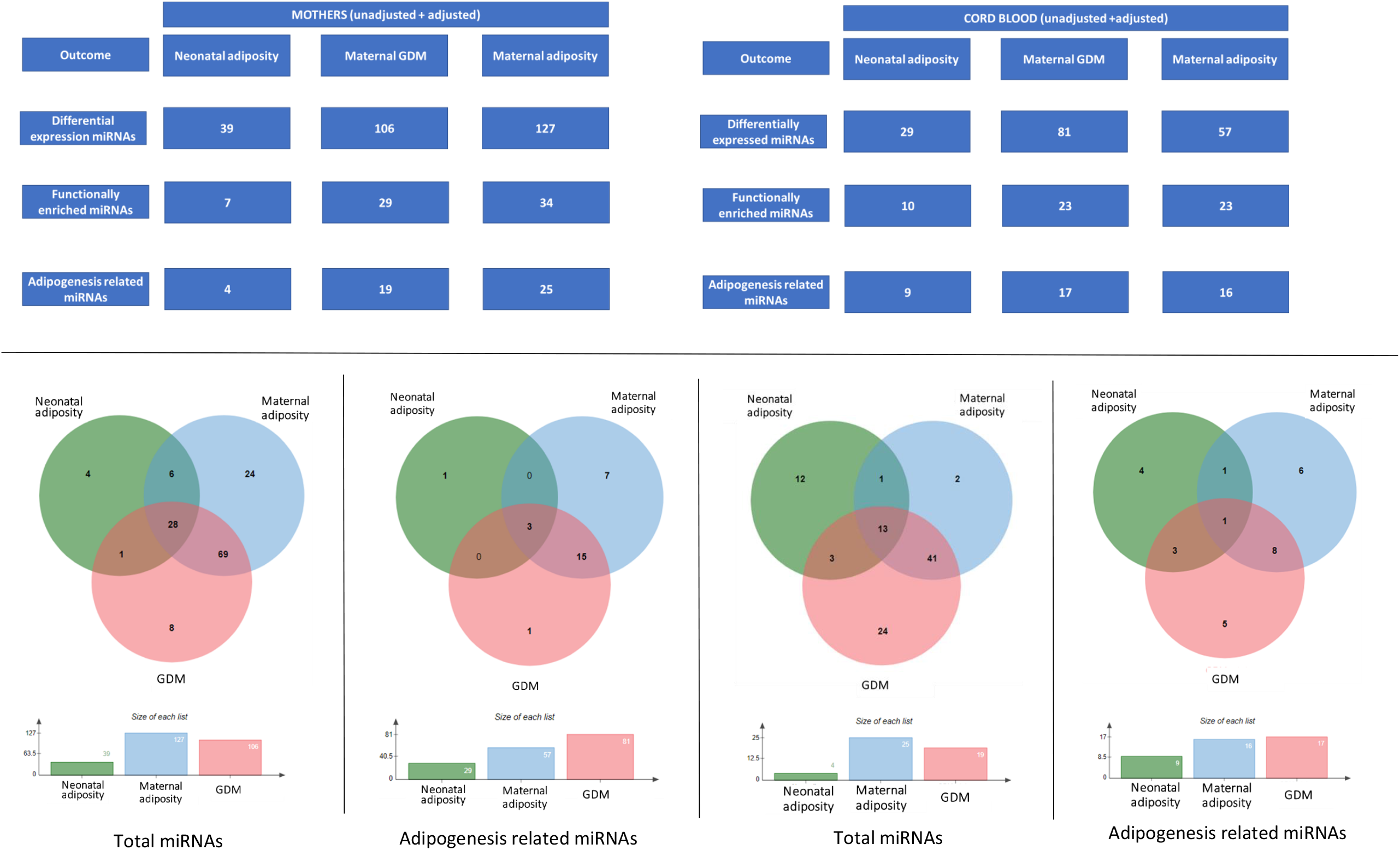
Comparison of differentially expressed miRNAs in the 3 analyses: The Venn diagram shows the number of differentially expressed miRNAs found common and exclusive to following 3 independent comparisons: 1, Neonatal adiposity (Lean Vs Adipose), 2. Maternal hyperglycemia during pregnancy (NGT Vs GDM), and 3. Maternal adiposity (Lean vs. Adipose)

## Discussion

There are few reports of ADsEV miRNAs in mother-neonate pairs. We investigated the association between ADsEV miRNAs in maternal and cord blood with neonatal body composition to understand their potential influence on neonatal adiposity. A large majority (∼85%) of ADsEV miRNAs in maternal and cord blood were common, and two-thirds of these were positively correlated suggesting a crosstalk between maternal adipose and neonatal tissue. Maternal adiposity and hyperglycemia, two major determinants of neonatal adiposity were both associated with differential expression of a relatively large number of ADsEV miRNAs in maternal and fetal circulation, a fifth of these were functionally related to adipogenesis. In the maternal circuit, these may represent adipose tissue dysfunction (i.e., adiposopathy) and have the potential to influence fetal growth and body composition.

Our primary analysis was to investigate maternal and cord blood miRNAs associated with neonatal adiposity. The tables 2 and 3 and the Sankey figure (Figure 4) show that the differentially expressed miRNAs were different in maternal and cord blood (except hsa-miR-212-3p which was common). Interestingly, they targeted the genes well-known to influence cellular signaling pathways related to adipogenesis. (FoxO, MAPK, Insulin, AGE-RAGE signaling, etc.) (figure 4). Literature survey revealed experimental evidence of the involvement of some of these miRNAs in aspects of adipogenesis. Maternal miRNAs hsa-miR-483-3p (FC= 3.99 in adipose group) and hsa-miR-212-3p (FC=0.282 in adipose group) have been associated with lipid accumulation and adipocyte differentiation in low-birth-weight adult humans^32^, and with increased subcutaneous adipose tissue inflammation^33^, respectively. In an earlier study we found that hsa-miR-212-3p is associated with increased risk of glucose intolerance in young adult PMNS female participants^34^. Fetal miRNAs hsa-miR-204-3p (FC=9.87 in adipose group) and hsa-miR-342-3p (FC=-0.098) have been shown to promote adipogenic differentiation of mesenchymal stem cells^35,36^ and hsa-miR-15a (FC= −0.912) to regulate intramuscular adipogenesis^37^. Given that neonates have a substantial proportion of brown adipose tissue (BAT) and that Indians have lower BAT compared to Europeans, we investigated if any of these DE miRNAs have originated in the BAT. Intriguingly, hsa-miR-92a-3p (FC= −0.36) and hsa-miR-181 (FC= 0.71 in adipose group) which are associated with brown fat activity^38,39^ are found to be differentially expressed only in cord blood, reflecting active brown fat activity in neonates. Together our results show a role for DE miRNAs in influencing maternal adipocyte function and fetal adipogenesis. We consider that our results provide an insight in the possible role of ADsEV miRNAs (maternal and fetal) in influencing neonatal adiposity and raise the possibility that they could act as maternal-fetal signaling system.

Growth and development of the fetus depends on coordinated timely expression of genes which is epigenetically regulated^40^. Body composition of the fetus is probably determined from very early pregnancy when the fate of the three germ layers is decided (gastrulation). Adipogenesis involves ‘commitment’ of mesenchymal stem cells to develop as pre-adipocytes which subsequently differentiate into adipocytes^41^. Several specific genes regulate these steps and the expression of these genes in turn is regulated by DNA methylation, histone modifications and specific miRNAs^42,43,44^. The role of DNA methylation in adipogenesis has been studied^45,46^ but there is little information on the other epigenetic mechanisms (i.e., miRNA regulation). Light microscopic studies in human abortuses demonstrate that pre-adipocyte to adipocyte differentiation happens between 19 to 23 weeks, after which period only adipocyte size increases by lipid accumulation^7^. Two major maternal determinants of these processes are maternal nutrition (both macro- and micro-nutrients) and her metabolism (diabetes and associated metabolic abnormalities)^47^. There is only scant information on how these two stimuli influence molecular adipogenesis in the baby, though insulin secretion by fetal pancreas is an important mechanism (Abigail Fouden). Maternal hyperglycemia promotes fetal hyperinsulinemia (Pedersen’s hypothesis)^48^, lipids and amino acids probably work similarly (Freinkel’s fuel mediated teratogenesis)^49^. In our novel approach we have investigated the possible involvement of maternal ADsEV miRNAs in influencing fetal adiposity and suggest that DE ADsEV miRNAs in both the maternal and neonatal circulation could influence neonatal adiposity through regulating various cellular signaling pathways which are well-known to be involved in different aspects of adipogenesis. Though our investigation does not allow us to confirm maternal origin of ADsEVs in fetal circulation, many studies have demonstrated maternal-fetal transfer of EVs.

Our study is perhaps the first of its kind to investigate association of maternal and fetal ADsEV miRNAs and neonatal adiposity. This is a novel approach to fetal growth and development and may provide useful leads into possible molecular mechanisms of maternal fetal communication. An additional strength is a reasonable number of mother-baby dyads in each group in a population which is characterized by adipose newborns (thin-fat Indian babies). One of the limitations is that the maternal ADsEV miRNAs were measured only at the time of delivery and thus after neonatal adiposity is established; we will investigate maternal ADsEVs in early pregnancy to expand our findings. It is also true that the current design is not provide conclusive proof of transfer of maternal ADsEVs to fetal circulation, and therefore our results are to be considered hypothesis generating and in need of further proof by experiments like HLA typing and matching of ADsEVs to track their origin. Moreover, as Indians are reported to have lower BAT compared to Europeans and it will be interesting to pursue adiposity trajectory and metabolic outcomes in these children in relation to cord blood ADsEVs. External validity of our findings will come only from additional studies in other populations.

In summary, we demonstrate differential expression of ADsEV miRNAs both in maternal and neonatal circulation in pregnancies with adipose neonates. These miRNAs targeted different aspects of adipose tissue function and adipogenesis, supporting a possible role in fetal adipogenesis. Studies in other populations and in early pregnancy will improve our understanding of maternal-fetal communication which influences fetal body composition.

## Data Availability

All data produced in the present study are available upon reasonable request to the authors.

## Abbreviations

PMNS: Pune Maternal Nutrition Study
PCS: Pune Children Study
SSF: Sum of skin folds
EV: Extracellular vesicles
ADsEV: Adipocyte derived small extracellular vesicles
GDM: Gestational diabetes mellitus
NGT: Normal glucose tolerant
FABP4: Fatty acid binding protein 4
DE: miRNA Differentially expressed microRNA

## Data availability

The demographic, phenotypic and raw microarray data is available with the authors and can be made available for a reasonable request following institutional clearances.

## Funding

The work was supported by InDiaGDM grant of the Department of Biotechnology, New Delhi, India (BT/IN/Denmark/02/CSY/2014) and National Institute of Health (NIH), the U.S government (R21HD094127-01).

## Acknowledgement

The authors thank Deepa Raut, Neelam Memane, Sayali Wadke and Rajashree Kamat for their help in processing cord blood samples and laboratory assays. Madhura Deshmukh for help in study coordination. Mrs. Pallavi Yajnik and Rasika Ladkat for administrative help. Dr. Leelavati Narlikar for her guidance in data analysis. Dr. Satyajit Rath for his advice in writing t discussion. We also thank the co-investigators of the InDiaGDM study Dr. Giriraj Chandak, Kalyanaraman Kumaran, Dr. Gundu Rao for their valuable contribution in conducting the InDiaGDM study.

## Author contributions

CSY designed the InDiaGDM study. CSY and HD contributed to managing the clinical cohort. DB, RKK, BH, ES, MB contributed to exosome extraction and protocol standardization. SR contributed to microRNA data generation. PSK, PT, MB, RKK, KA, MG contributed to the data analysis. PSK, MB, CSY, RF contributed to the manuscript writing. CSY and RF contributed to discussion and interpretation of the results. All authors read and approved the manuscript.

---

**Supplementary figure 1: STROBE flow diagram for the selection of participants**

**Supplementary figure 2: STROBE Flow diagram for miRNA analysis**

**Supplementary figure 3: Comparison of average z-score of miRNA expression profile in maternal and cord blood samples between lean Vs adipose groups:** The boxplots highlight that (A) the average z-score of all miRNA expression was lower in mothers of adipose neonates (red color) as compared to mothers of lean neonates (green color) (p=0.053). This pattern was not observed in the cord blood (B) of adipose and lean neonates (p=0.645). The z-scores were calculated as per the formula:

Z-score = average (expression of miRNA in each sample - median expression of that miRNA)/ Std. dev(miRNA))

## Notes

### Competing Interest Statement

The authors have declared no competing interest.

### Funding Statement

The study was funded by InDiaGDM grant of the Department of Biotechnology, New Delhi, India (BT/IN/Denmark/02/CSY/2014) and National Institute of Health (NIH), the U.S government (R21HD094127-01).

### Author Declarations

Ethics committee of King Edward Memorial Hospital Research Centre, Pune, India gave ethical approval for this work

