## Supplementary figures and images for "MicroRNAs in adipocyte-derived extracellular vesicles in maternal and cord blood are related to neonatal adiposity"

### Supplementary figure 1

**Supplementary figure 1:** STROBE flow diagram for the selection of participants

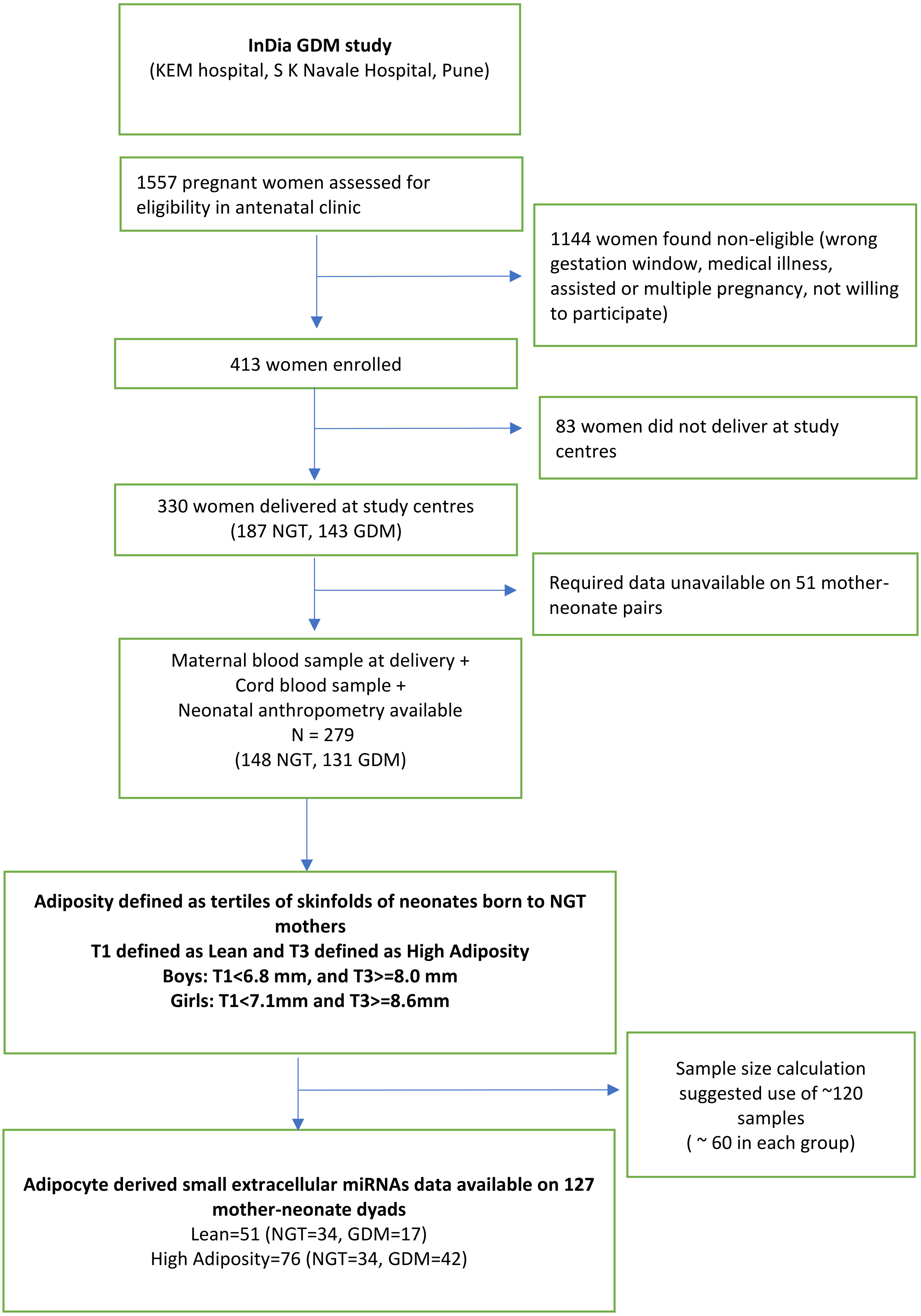

### Supplementary figure 2

#### Supplementary figure 2: STROBE Flow diagram for miRNA analysis

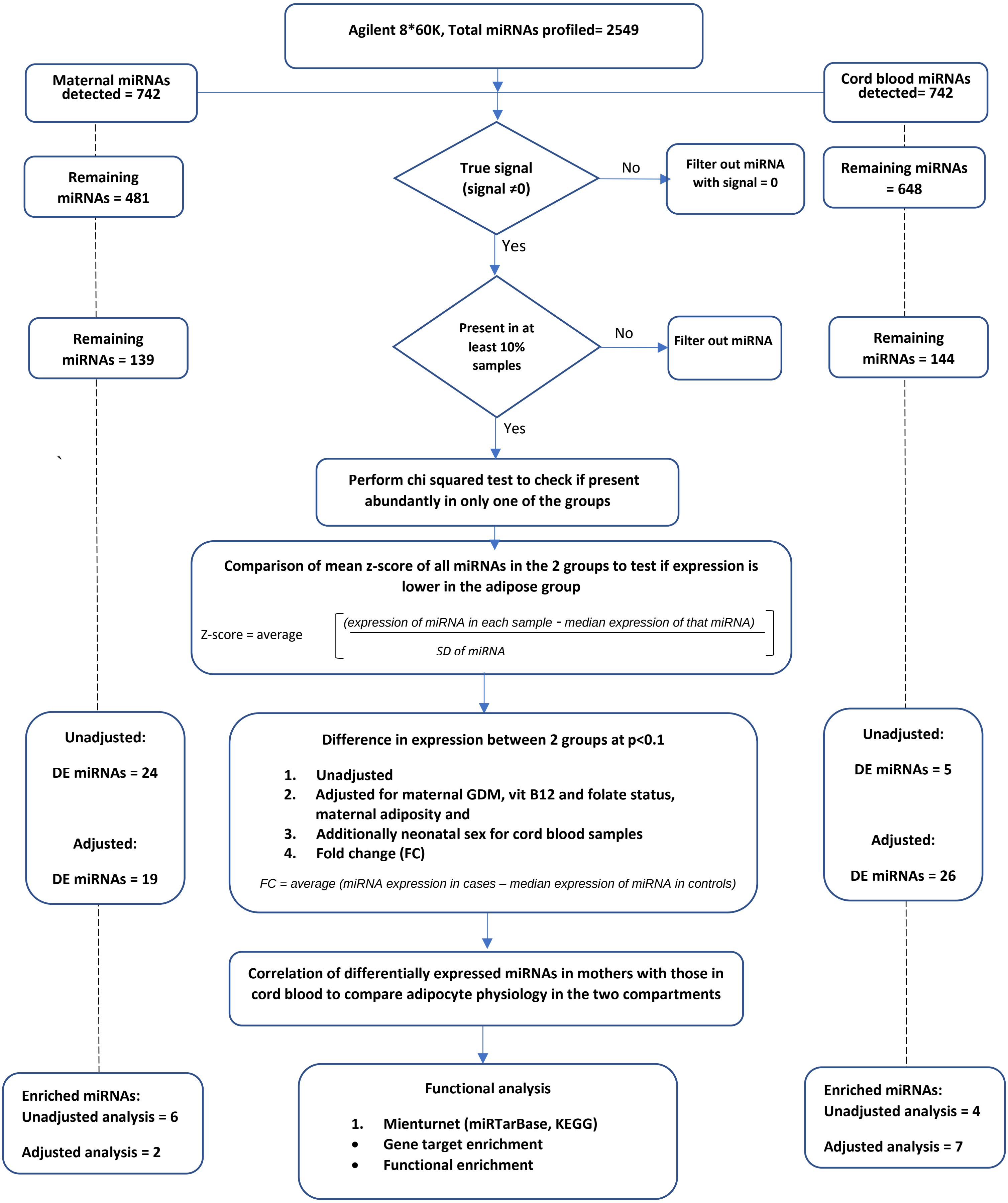
