## Supplementary table 1 for "MicroRNAs in adipocyte-derived extracellular vesicles in maternal and cord blood are related to neonatal adiposity"

**Supplementary table 1. Laboratory methods**

| **Analyte** | **Test** | **Instrument** | **Comment** |
| --- | --- | --- | --- |
| **Complete Blood Count** | Whole blood collected and analysed on the day of visit | Beckman Coulter analyser (AC.T diffTM Analyzer, Miami, Florida, USA). | - |
| **Glucose** | Glucose oxidase (GOD)- Peroxidase) POD. | Spectrum; Abbott, (Irving, TX). | ·         Between-batch coefficients of variation for all the assays were <3% in the normal range |
| **Insulin** | ELISA (Mercodia AB, SE-754 50, Uppsala, Sweden). | Mercodia AB , Uppsala, Sweden. | ·         Intra- and inter-assay CV <7% |
| **Vitamin B12** | Microbiological assay on plasma using a colistin sulfate-resistant strain of L. Leichmanii | - | ·         This is our standard research assay. |
|  |  |  | ·         Sensitivity: 50 pmol/L |
|  |  |  | ·         CV: <8% |
|  |  |  | ·         Antibiotic treatment interferes with this assay. |
|  | Electro Chemiluminescence Immuno Assay (ECLIA) on plasma | Cobas e-411 analyzer, ROCHE diagnostics GmbH, Sandhofer Strasse 116, Mannheim, Germany | ·         We used this assay because women undergoing caesarean section received antibiotics before delivery which interferes with microbial assay. |
|  |  |  | ·         Correlation coefficient with Microbiological assay r = 0.958 by paired t-test |
|  |  |  | ·         Sensitivity: 62 pmol/L |
|  |  |  | ·         CV: 7% |
| **Holo-TC (Holo-transcobalamin)** | Chemiluminescent microparticle immunoassay (CMIA) on plasma, using anti-holotranscobalamin coated paramagnetic microparticles | ARCHITECT (ABBOTT-GmbH &amp; Co. KG Max-Planck-Ring2 65205 Wiesbaden Germany +49-6122-580) | ·         Sensitivity: <= 5.0 pmol/L |
|  |  |  | ·         CV: <= 8.5 % |
| **Folate** | Microbiological assay on plasma using a chloramphenicol-resistant strain of L. Casei | - | ·         Sensitivity: 3nmol/L |
|  |  |  | ·         CV: <8% |
| **Total homocysteine** | HPLC on plasma | PerkinElmer 200 Series, PerkinElmer, Shelton, CT, USA, using fluorescence detector | ·         Sensitivity: 3 µmol/L |
|  |  |  | ·         CV: <4% |
| **Cholesterol** | Measured using standard enzymatic kits for oxidase peroxidase reaction | Spectrum; Abbott, (Irving, TX). | ·         Between-batch coefficients of variation for all the assays were <3% in the normal range |
| **Triglycerides** |  |  |  |
| **HDL** |  |  |  |
