## Supplementary table 2 for "MicroRNAs in adipocyte-derived extracellular vesicles in maternal and cord blood are related to neonatal adiposity"

**Supplementary table 2. Comparison of demographic characteristics between the participants included and those excluded**

| MOTHER DATA AT DIAGNOSIS OF GDM (bias statement) | | | | | |
| --- | --- | --- | --- | --- | --- |
|  | EXCLUDED | | INCLUDED | | P - VALUE |
|  | N | MEDIAN (LQ , UQ) | N | MEDIAN (LQ , UQ) |  |
| MOTHER | | | | | |
| Age (yrs) | 148 | 26.9  ( 24.4 , 30.2 ) | 125 | 27.2  ( 24.7 , 30.7 ) | 0.652 |
| Height (cm) | 150 | 154.4 ( 5.5 ) | 126 | 154.9 ( 5.7 ) | 0.462 |
| Weight (kg) | 150 | 61.8  ( 54.5 , 67.9 ) | 126 | 64.9  ( 56.8 , 72.3 ) | 0.049 |
| BMI (kg/m²) | 148 | 25.8  ( 23.2 , 29.1 ) | 126 | 27.1  ( 23.5 , 30.1 ) | 0.093 |
| Subscapular Skinfold (mm) | 145 | 36.3  ( 28.2 , 45 ) | 125 | 38  ( 28.6 , 45.9 ) | 0.675 |
| Fasting Glucose (mg%) | 148 | 80  ( 74 , 89 ) | 122 | 82  ( 76 , 89.8 ) | 0.164 |
| Fasting Insulin (IU/L) | 143 | 8.7  ( 5.8 , 12.9 ) | 117 | 9  ( 6.7 , 14.1 ) | 0.372 |
| iHoma Sensitivity | 143 | 103.6  ( 72.6 , 157.4 ) | 117 | 101.5  ( 66.2 , 137.5 ) | 0.352 |
| iHoma Beta | 143 | 122.9  ( 93.5 , 151.4 ) | 117 | 118.3  ( 95.5 , 147.6 ) | 0.706 |
| iDI (*10³) | 143 | 13085  ( 9720.5 , 16314.3 ) | 117 | 12488.8  ( 9149 , 15240.2 ) | 0.174 |
| Cholesterol (mg%) | 145 | 199.2 ( 43.9 ) | 120 | 197.6 ( 41.3 ) | 0.548 |
| Triglycerides (mg%) | 145 | 150  ( 127 , 184 ) | 120 | 144  ( 115 , 200.2 ) | 0.631 |
| HDL (mg%) | 145 | 57.9 ( 12.2 ) | 120 | 57.3 ( 13.7 ) | 0.59 |
| Vitamin B12 (pM) | 149 | 211  ( 153 , 280 ) | 124 | 213.5  ( 159.8 , 280 ) | 0.64 |
| OFFSPRING | | | | | |
| Gender | **N** | **Male=73, Female=79** | **N** | **Male=61, Female=66** | P - VALUE |
| Gestation Age at Delivery (days) | 149 | 225  ( 202 , 244 ) | 126 | 226.5  ( 212.2 , 244.8 ) | 0.452 |
| Birth Weight (kg) | 151 | 2845.9 ( 389.5 ) | 127 | 2829.4 ( 446.3 ) | 0.962 |
| Birth Length (cm) | 152 | 48.7  ( 47.5 , 49.8 ) | 127 | 48.3  ( 47.2 , 49.5 ) | 0.249 |

**Note:** Data are presented as mean(SD) and p-values are calculated by Student’s T-test if data is normally distributed whereas median (25^th^, 75^th^ centile) and p-values calculated by Mann-Whitney are presented if data is not normally distributed.

BMI: Body Mass Index; HOMA: Homeostatic Model Assessment models, DI: Disposition index. SD: Standard deviation.
