## Supplementary figure 3 for "MicroRNAs in adipocyte-derived extracellular vesicles in maternal and cord blood are related to neonatal adiposity"

**Supplementary figure 3. Comparison of average z−score of miRNA expression profile in maternal and cord blood samples between lean Vs adipose groups**

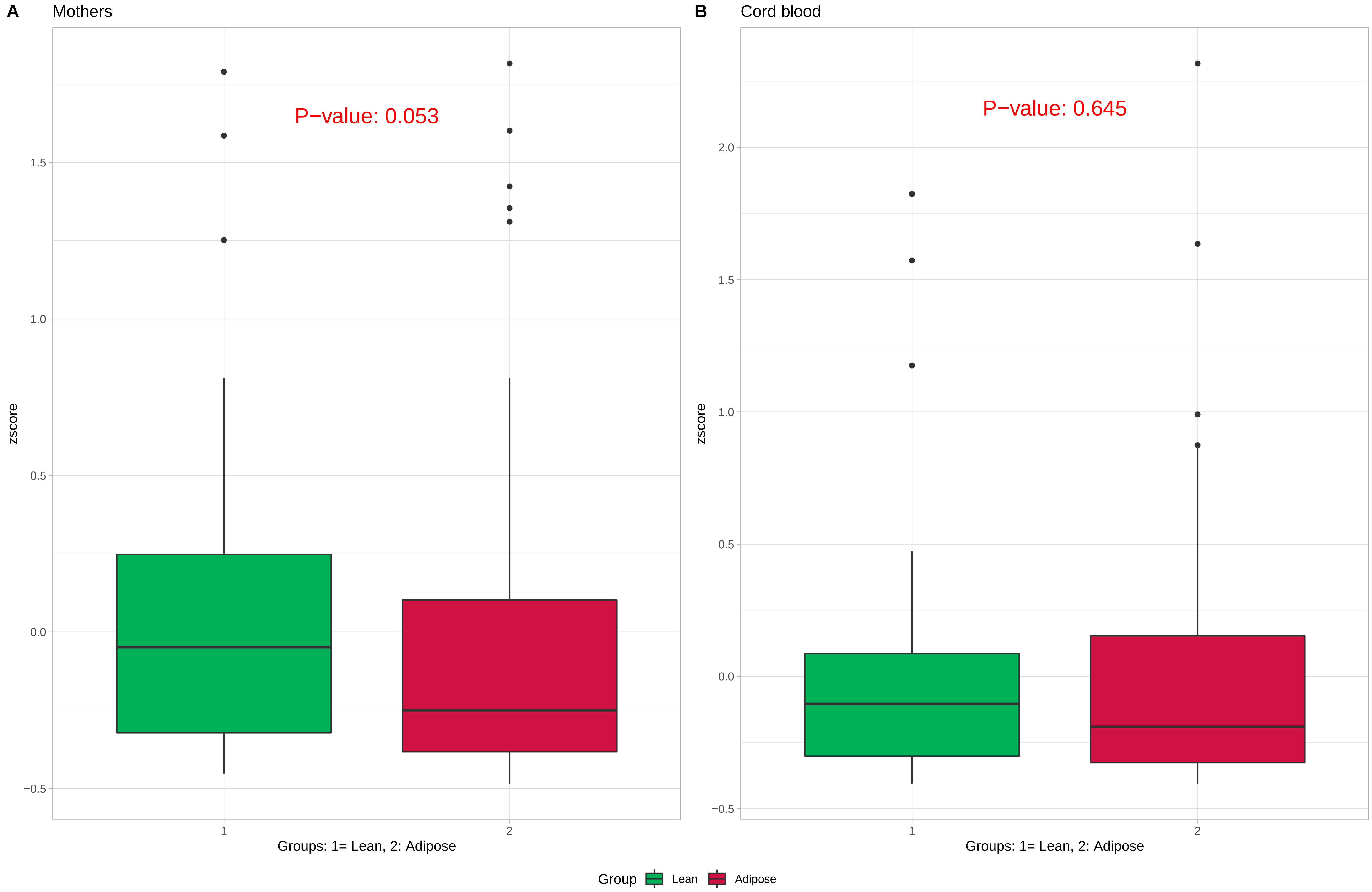
